# Replicating extensive brain structural heterogeneity in individuals with schizophrenia and bipolar disorder

**DOI:** 10.1101/2020.05.08.20095091

**Authors:** Thomas Wolfers, Jaroslav Rokicki, Dag Alnæs, Pierre Berthet, Ingrid Agartz, Seyed Mostafa Kia, Tobias Kaufmann, Mariam Zabihi, Torgeir Moberget, Ingrid Melle, Christian F Beckmann, Ole A Andreassen, Andre F Marquand, Lars T Westlye

## Abstract

Identifying brain processes involved in the risk and development of mental disorders is a major aim. We recently reported substantial inter-individual heterogeneity in brain structural aberrations among patients with schizophrenia and bipolar disorder. Estimating the normative range of voxel-based morphometry (VBM) data among healthy individuals using a gaussian process regression (GPR) enables us to map individual deviations from the healthy range in unseen datasets. Here we aim to replicate our previous results in two independent samples of patients with schizophrenia (n1=94; n2=105), bipolar disorder (n1=116; n2=61) and healthy individuals (n1=400; n2=312). In line with previous findings with exception of the cerebellum our results revealed robust group level differences between patients and healthy individuals, yet only a small proportion of patients with schizophrenia or bipolar disorder exhibited extreme negative deviations from normality in the same brain regions. These direct replications support that group level-differences in brain structure disguise considerable individual differences in brain aberrations, with important implications for the interpretation and generalization of group-level brain imaging findings to the individual with a mental disorder.

## INTRODUCTION

Recently, the degree of inter-individual heterogeneity in brain structure was found to be considerably larger than previously anticipated for both schizophrenia and bipolar disorder [Wolfers et al., 2018]. As expected, based on the substantial body of literature reporting results from case-control comparisons, patients with schizophrenia or bipolar disorder show evidence of group level deviations from a normative trajectory in brain structure. However, applying normative modeling [Marquand et al., 2016; Marquand et al., 2019] to chart variation in brain anatomy across individual patients showed highly idiosyncratic patterns of deviation, suggesting that such group effects are inaccurate reflections of the brain aberrations found at the individual level [Wolfers et al., 2018]. Of note, a similar high level of heterogeneity has recently also been observed in attention-deficit/ hyperactivity disorder [Wolfers et al., 2019] and autism spectrum disorder [Zabihi et al., 2019].

Given the existing literature on reproducible group-level differences in brain structure between cases and controls [Van Erp et al., 2016; Moberget et al., 2017], our initial findings of substantial heterogeneity within disorders demonstrated that moving beyond the study of group differences is highly beneficial to understand variability within clinical cohorts and may be required to make inferences at the level of the individual. Due to these important implications, we here report an attempt to replicate our initial findings in two independent samples acquired on different scanners following an identical analytical procedure as in our previous discovery study.

## METHODS

### Participants

Table 1 summarizes the demographic and clinical information of the replication samples and the sample used in the discovery publication [Wolfers et al., 2018]. For replication sample 1 we included 94 patients with a schizophrenia diagnosis, 116 patients with a bipolar disorder diagnosis and 400 healthy individuals. As the replication sample 2 we included 312 healthy individuals, 105 with schizophrenia diagnosis and 61 with bipolar diagnosis. As the discovery sample we selected 256 healthy individuals, 163 patients with schizophrenia and 190 with bipolar disorder. All participants were recruited from the same population and catchment area but there was no overlap between the discovery and replication samples. All participants were recruited as part of the Thematically Organized Psychosis (TOP) study, approved by the Regional Committee for Medical Research Ethics and the Norwegian Data Inspectorate [Doan et al., 2017]. The two replication samples were selected from the TOP-database on the 25^th^ of September 2019. Patients were recruited from in- and out-patient clinics in the Oslo area, understood and spoke a Scandinavian language, had no history of severe head trauma, and had an IQ above 70. Patients were assessed by trained physicians or clinical psychologists. Psychiatric diagnosis was established using the Structured Clinical Interview for DSM-IV Axis I Disorders (SCID). Symptoms were assessed using PANSS [Kay et al., 1987]. We used the positive, negative and global summary scores of the PANSS which were combined to the total summary score. Healthy individuals were randomly sampled from national registries and neither they nor their relatives had a psychiatric or alcohol/substance use disorder or cannabis use during the last 3 months. Written informed consent was obtained from all participants.

**Table 1:** Demographics.

| Demographics | Replication 2<sup>#</sup> |  |  | Replication 1<sup>#</sup> |  |  | Discovery |  |  |
| --- | --- | --- | --- | --- | --- | --- | --- | --- | --- |
|  | Healthy | BP | SZ | Healthy | BP | SZ | Healthy | BP | SZ |
| N | 312 | 61 | 105 | 400 | 116 | 94 | 256 | 190 | 163 |
| Male (%) | 58.01% | 45.90% | 64.76% | 50.25% | 35.34% | 57.44% | 54.70% | 41.80% | 64.40% |
| Age | 30 ± | 31 ± | 27 ± | 34 ± | 31 ± | 28 ± | 34 ± | 34 ± | 31 ± |
| (mean ± std) | 8.0 | 11.8 | 8.8 | 11.3 | 10.8 | 9.2 | 9.5 | 11.3 | 8.7 |
| Years of education | 14.3± | 13.8± | 11.9± | 14.4± | 13.8± | 12.7± | 14.0 ± | 13.6 ± | 12.9 ± |
| (mean ± std) | 2.3 | 2.1 | 2.1 | 2.4 | 2.2 | 2.3 | 2.3 | 2.3 | 2.6 |
| **Symptom scores\*** |  |  |  |  |  |  |  |  |  |
| PANSS global | na | 24.2± | 30.9± | na | 25.5 ± | 30.6 ± | na | 25.4 ± | 32.1 ± |
| (mean ± std) |  | 4.8 | 8.8 |  | 5.3 | 7.6 |  | 5.7 | 8.6 |
| PANSS negative | na | 9.3± | 16.2± | na | 9.6 ± | 15.7 ± | na | 10.1 ± | 15.8 ± |
| (mean ± std) |  | 3.1 | 6.2 |  | 5.3 | 6.4 |  | 3.5 | 6.3 |
| PANSS positive | na | 9.0± | 14.5± | na | 9.3 ± | 13.4 ± | na | 10.0 ± | 15.1 ± |
| (mean ± std) |  | 2.7 | 5.7 |  | 3.2 | 4.3 |  | 3.6 | 5.5 |
| PANSS total | na | 42.6± | 61.6± | na | 44.5 ± | 59.8 ± | na | 45.5 ± | 63.1 ± |
| (mean ± std) |  | 7.9 | 18.2 |  | 9.3 | 15.9 |  | 10.0 | 16.8 |

Abbreviations: na, not applicable; PANSS, Positive and Negative Syndrome Scale. BP, bipolar disorder. SZ, schizophrenia.
\* Symptom scores have been assessed using PANSS which is a standard clinical instrument for the quantification of positive and negative psychotic symptoms.

### MRI acquisition

*Discovery:* Structural scans were obtained on 1.5 Tesla Siemens MAGNETOM Sonata scanner at Oslo University Hospital using a standard 32-channel head coil. T1-weighted images were acquired using a MPRAGE sequence with the following parameters: repetition time (TR) = 2730ms, echo time (TE) = 3.93ms, flip angle (FA) = 7°. *Replication 1:* Structural scans were obtained on 3 Tesla GE 750 Discovery scanner at Oslo University Hospital using a standard 32-channel head coil. T1-weighted images were acquired using a BRAVO sequence with the following parameters: repetition time (TR) = 8.16ms, echo time (TE) = 3.18ms, flip angle (FA) = 12°. *Replication 2:* Structural scans were obtained on 3 Tesla GE Signa HDxT at Oslo University Hospital one subset with HNS coil the other subset with 8HRBRAIN coil. T1-weighted images were acquired using the following parameters: repetition time (TR) = 7.8ms, echo time (TE) = 2.956ms, and flip angle (FA) = 12.

### Estimation of gray matter volume

In the same way as in our previous study, raw T1-weighted MRI volumes were processed using the computational analysis toolbox version 12 (CAT12; http://www.neuro.uni-jena.de/software/), based on statistical parametric mapping version 12 (SPM12). Images were segmented, normalized, and bias-field-corrected using VBM-SPM12 (http://www.fil.ion.ucl.ac.uk/spm, London, UK) [Ashburner and Friston, 2000; Ashburner and Friston, 2003], yielding images containing gray and white matter segments. Prior to the estimation of the normative models, all gray and white matter volumes were smoothed with an 8 mm FWHM Gaussian smoothing kernel and we restricted our analyses to voxels included in the gray matter mask constructed for the discovery study.

### Normative modeling

As in our previous article, we estimated the normative model using Gaussian Process Regression (GPR) to predict VBM based regional gray matter volumes across the brain from age and sex. To avoid overfitting of the normative models, it is crucial to estimate predictive performance out of sample. Therefore, we estimated the normative range for this model in healthy individuals under 10-fold cross-validation, and then applied one model across all healthy individuals to patients with schizophrenia and bipolar disorder. GPR yields coherent measures of predictive confidence in addition to point estimates. This is important in normative modelling as we need this uncertainty measure to quantify the deviation of each patient from the group mean at each brain locus. Thus, we are able to statistically quantify deviations from the normative model with regional specificity, by computing a Z-score for each voxel reflecting the difference between the predicted and the observed gray matter volume normalized by the uncertainty of the prediction [Marquand et al., 2016].

In line with our previous article, we thresholded the individual normative probability maps at p<.005 (i.e. |Z|>2.6) and extreme positive and extreme negative deviations from the normative model were defined based on this threshold. All extreme deviations were combined into scores representing the percentage of extreme positively and negatively deviating voxels for each participant, relative to the total number of voxels in the brain mask. We tested for associations between diagnosis and those scores using a non-parametric test corrected for multiple comparisons using the Bonferroni-Holm method [Holm, 1979] as well as an association with PANSS scores. We repeated these analyses using different thresholds p<0.05 (i.e. |Z|>1.96) as well as p<0.001 (i.e. |Z|>3.1) and also modeled extreme deviations using extreme value statistics [Fisher and Tippett, 1928]. This is based on the notion that the expected maximum of any random variable converges to an extreme value distribution. Therefore, we estimated a maximum deviation for each subject by taking a trimmed mean of 1% of the top absolute deviations for each subject across all vertices and fit an extreme value distribution to these deviations. Thus in addition to our previous work we checked whether our results remain consistent independent of the thresholding procedure of the normative probability maps that we have introduced in different publications [Marquand et al., 2016; Wolfers et al., 2018; Wolfers et al., 2019; Zabihi et al., 2019]. To assess the spatial extent of those extreme deviations, we created individualized maps and calculated the voxel-wise overlap between individuals from the same groups first by replicating the exact procedure of the discovery study then by introducing different thresholds to check consistency. In the main text we report this overlap for healthy individuals, and people diagnosed with schizophrenia and bipolar disorder. All analyses were performed in python3.6 (www.python.org) and scripts are available on GitHub (https://github.com/RindKind/). Also, in line with our previous article, we fed the normative probability maps into PALM [Winkler et al., 2015] to test for mean differences between groups by means of a general linear model framework and permutation-based inference.

## RESULTS

### Normative modeling

Figure 1 shows the spatial representation of the voxel-wise normative model, characterized by widespread gray matter decreases from age 20 to 70, with most pronounced age-differences in frontal areas. We depict models for discovery and replication studies separately in this figure. Further, we could show that the models performed well across the whole brain by plotting the correlation of predicted and observed values under 10-fold cross validation. This is depicted in Supplementary Figure 1.

**Figure 1:**
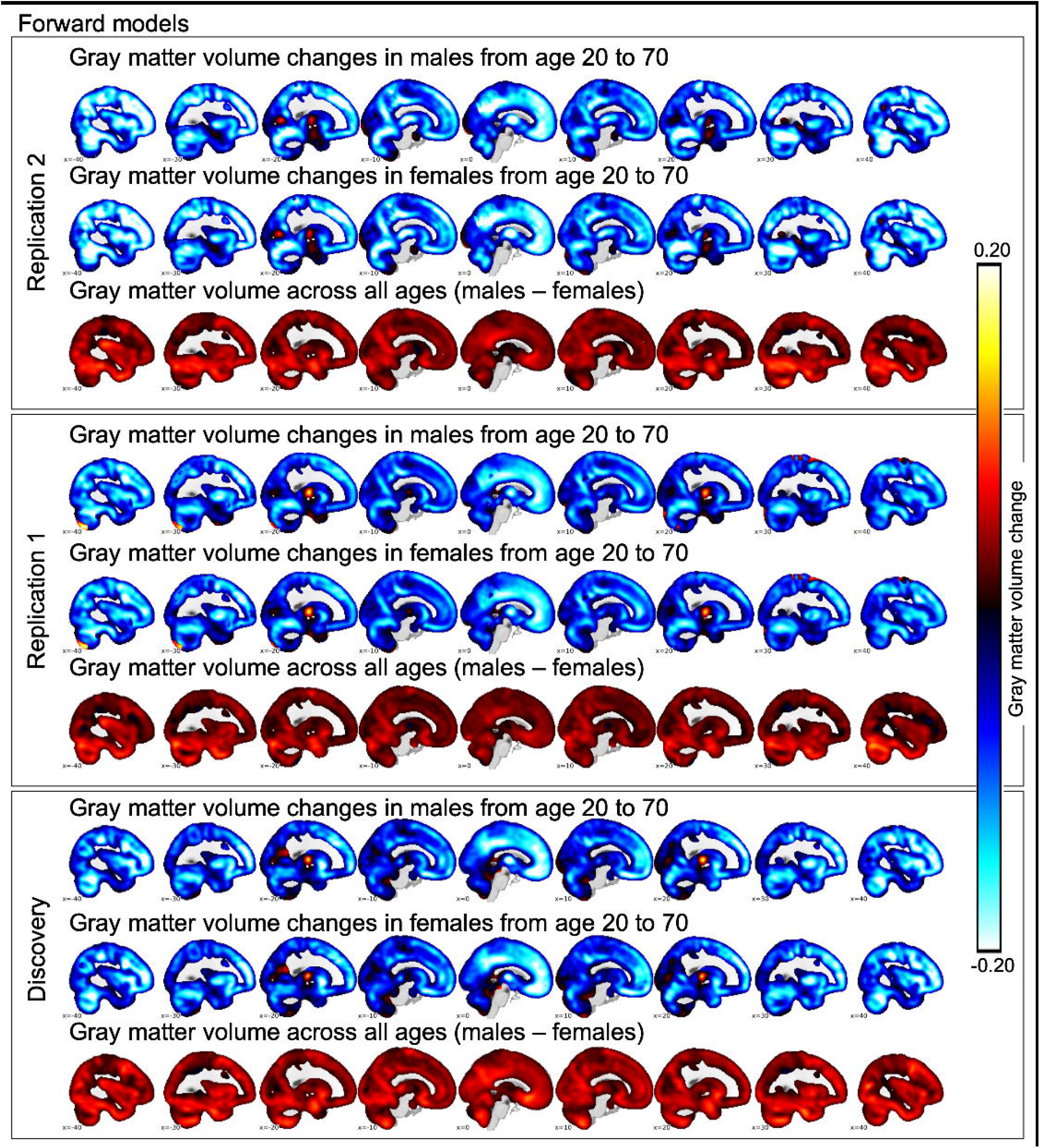
We depict the slope of a linear approximation of the normative model for males (first row in each panel) and females (second row in each panel) as well as the difference between males and females across the entire age range from 20-70 years (third row in each panel). In the lower panel we depict results based on the data reported in Wolfers et al. 2018, Jama Psychiatry. In the upper two panels we depict two replications. Note: These approximations are based on the forward model of the estimated normative models.

### Group comparisons

Figure 2 shows the result from pairwise group comparisons, corrected for multiple comparisons using permutation testing in PALM. In gray matter, patients with schizophrenia show stronger mean negative deviations than healthy individuals in frontal, temporal, and cerebellar regions; mean deviations are also more negative than in patients with bipolar disorder and localized primarily in frontal brain regions (Supplementary Figure 2). These results replicate well across the three samples. However, for bipolar disorder the replication is not as strong showing differences from healthy controls in the cerebellum only in the discovery sample but not in the two replication samples. This may be due to a lower sample size of the bipolar and schizophrenia groups in both replication samples. The differences between patients with schizophrenia diagnosis and healthy individuals are very robust.

**Figure 2:**
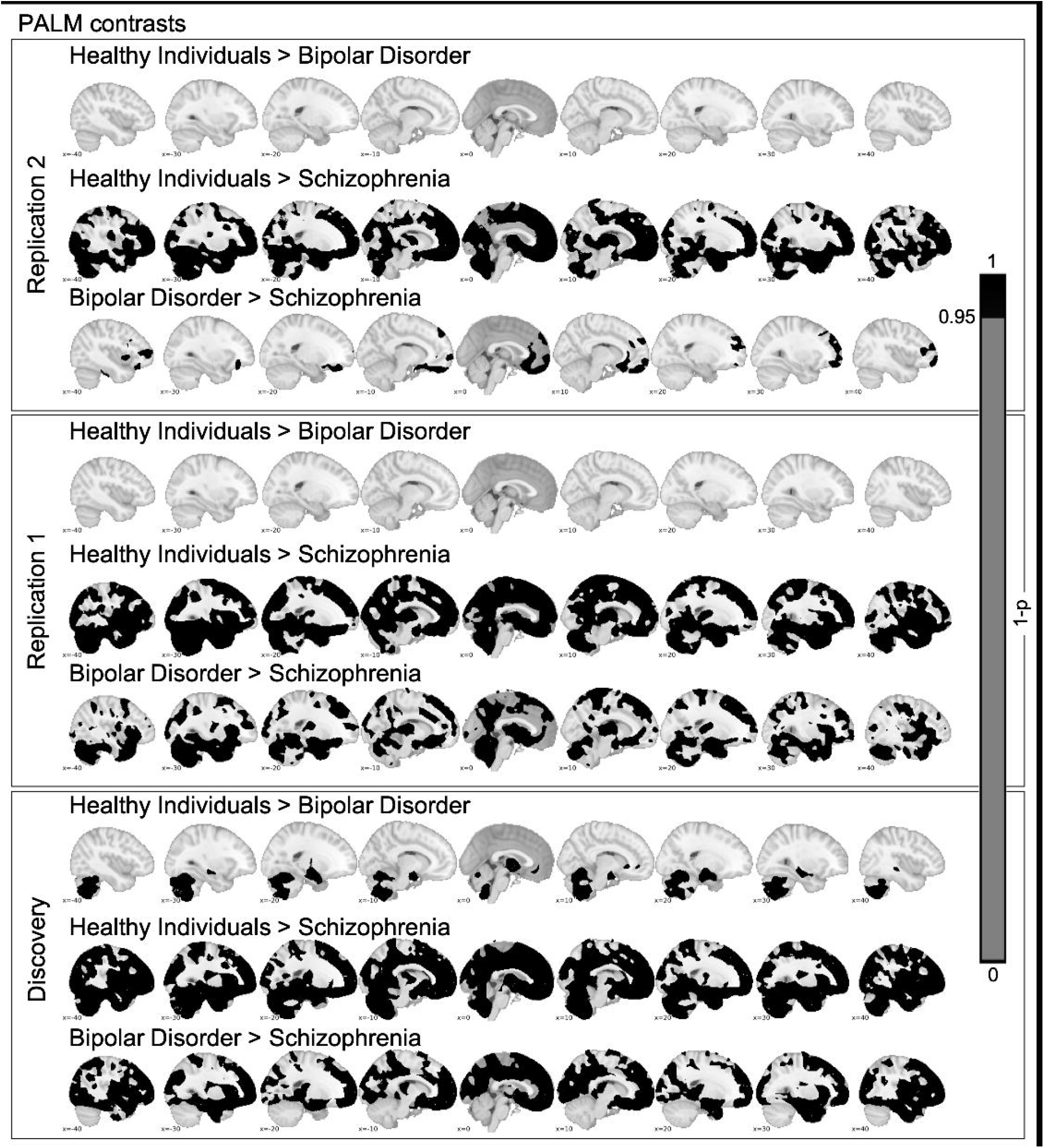
We depict the contrast between healthy individuals, bipolar disorder and schizophrenia. In the lower panel we depict results based on the data reported in Wolfers et al. 2018, Jama Psychiatry. In the upper two panels we depict two replications. For the PALM derived mean Z-scores see Supplementary Figure 2. Note: We report one subtracted by multiple comparison corrected p-values.

### Spatial distribution and statistical analyses of extreme deviations from normality

In line with our discovery study, in replication study 1 patients with schizophrenia show a higher percentage of extreme negative deviations across the brain (0.64 ± 1.15% of all voxels) compared to healthy individuals (0.16 ± 0.44%, Mann-Whitney p<0.001) and individuals with bipolar disorder (0.14 ± 0.34%, Mann-Whitney p<0.001, Table 2). The percentage of extreme positive deviations across participants and groups show that healthy individuals (1.16 ± 1.99%) differed from patients with schizophrenia (0.60 ± 0.90%; Mann-Whitney p <0.001) and from individuals with bipolar disorder (0.88 ± 1.44%; Mann-Whitney p <0.05). In replications study 2, patients with schizophrenia show a higher percentage of extreme negative deviations across the brain (1.09 ± 4.09% of all voxels) compared to healthy individuals (0.22 ± 0.53%, Mann-Whitney p<0.001) and individuals with bipolar disorder (0.35 ± 1.11%, Mann-Whitney p<0.001, Table 2). The percentage of extreme positive deviations across participants and groups show that healthy individuals (1.03 ± 2.06%) differed from patients with schizophrenia (0.85 ± 1.59%; Mann-Whitney p <0.001) and from individuals with bipolar disorder (0.98 ± 2.59%; Mann-Whitney p <0.05). This is an exact replication of the previous study (Table 2). Extreme negative deviations are on average 3.91, 4.00 and 4.95 times more prevalent in individuals with schizophrenia than in healthy controls across discovery and replication samples 1/2, respectively (Supplementary Figure 3/4). All these results replicate for different Z-thresholds on the normative probability maps (Supplementary Table 1) and also remain consistent with an estimate based on extreme value statistics (Supplementary Table 1). Further we show that extreme negative deviations correlate significantly with symptom scores as measured by the PANSS across all samples, (discovery sample: r=0.241, p<0.001; replication sample 1: r=0.157, p<0.05; replication sample 2: r=0.190 p<0.05; Table2). This shows that increasing number of symptoms is associated with more extreme negative deviations. This effect was only found across patient groups not within patient groups (Supplementary Table 2), which may be due to lower power in individual groups or could potentially reflect a group difference rather than a dimensional effect across groups. Further, we could show an association of extreme negative deviations with the age of onset of both schizophrenia and bipolar disorder but not with other clinical characteristics, the duration of medication or lifetime episodes of psychotic, depressive, manic, or hypomanic events (Supplementary Table 3/4).

**Table 2:** Extreme deviations (|Z|>2.6)

|  | Replication 2 |  |  | Replication 1 |  |  | Discovery |  |  |
| --- | --- | --- | --- | --- | --- | --- | --- | --- | --- |
|  | Healthy | BP | SZ | Healthy | BP | SZ | Healthy | BP | SZ |
| **Case-control** |  |  |  |  |  |  |  |  |  |
| **Extreme negative** | 0.22 +-<br>0.53% | 0.35 +-<br>1.11% | 1.09 +-<br>4.09% | 0.16 +-<br>0.44% | 0.14 +-<br>0.34% | 0.64 +-<br>1.15% | 0.23 +-<br>0.78% | 0.24 +-<br>0.48% | 0.90 +-<br>2.15% |
| **(mean, std)** |  |  |  |  |  |  |  |  |  |
| **Significance** | HC = BD<br>HC < SZ ($p < 0.001$)<br>BP < SZ ($p < 0.01$) | | | HC = BD<br>HC < SZ ($p < 0.001$)<br>BP < SZ ($p < 0.001$) | | | HC = BD<br>HC < SZ ($p < 0.001$)<br>BP < SZ ($p < 0.001$) | | |
| **Extreme positive** | 1.03 +-<br>2.06% | 0.98 +-<br>2.59% | 0.85 +-<br>1.59% | 1.16 +-<br>1.99% | 0.88 +-<br>1.44% | 0.60 +-<br>0.90% | 1.08 +-<br>1.75% | 0.79 +-<br>1.25% | 0.78 +-<br>1.50% |
| **(mean, std)** |  |  |  |  |  |  |  |  |  |
| **Significance** | HC = BD<br>HC > SZ ($p < 0.05$)<br>BP = SZ | | | HC = BD<br>HC > SZ ($p < 0.001$)<br>BP > SZ ($p < 0.05$) | | | HC = BD<br>HC > SZ ($p < 0.001$)<br>BP > SZ ($p < 0.05$) | | |
| **Dimensional** | BP & SZ |  |  | BP & SZ |  |  | BP & SZ |  |  |
| **Extreme negative** | r=0.190 |  |  | r=0.157 |  |  | r=0.241 |  |  |
| **PANSS total\*** | p<0.05 |  |  | p<0.05 |  |  | p<0.001 |  |  |
| **Extreme positive** | r=0.071 |  |  | r=-0.072 |  |  | r=0.035 |  |  |
| **PANSS total\*** | p>0.05 |  |  | p>0.05 |  |  | p>0.05 |  |  |

Abbreviations: PANSS, Positive and Negative Syndrome Scale. BP, bipolar disorder. SZ, schizophrenia.
\* Symptom scores have been assessed using PANSS which is a standard clinical instrument for the quantification of positive and negative psychotic symptoms.

Extreme negative deviations in people with schizophrenia were most pronounced in temporal, medial frontal and posterior cingulate regions (Figure 3). In patients with bipolar disorder the overlap was strongest in the thalamus. In line with our previous findings the overlap of extreme negative (Figure 3) and positive deviations from normality (Supplementary Figure 5) is sparse across individuals with the same diagnosis, with peak voxels showing extreme negative overlap in 2.75% healthy individuals, 5.17% for individuals with bipolar disorder and 9.57% for schizophrenia in replication sample 1. In replication sample 2, the peak voxel shows extreme negative overlap in 3.52% of the healthy individuals, 8.19% of the individuals with bipolar disorder and 9.52% for individuals with a schizophrenia diagnosis. In expectation this overlap increased when we applied a lower threshold |Z|> 1.96 (Supplementary Figure 6) but was still sparse and decreased when we utilized a higher threshold |Z|> 3.1 (Supplementary Figure 7) or and FDR threshold equal to 0.05 (Supplementary Figure 8). Independent of the threshold the findings of the discovery study were replicated across two samples. Interestingly when we stratified for sex the overlap of extreme negative deviations was stronger in males, which was consistent across samples and true for both disorders (Supplementary Figure 9).

**Figure 3:**
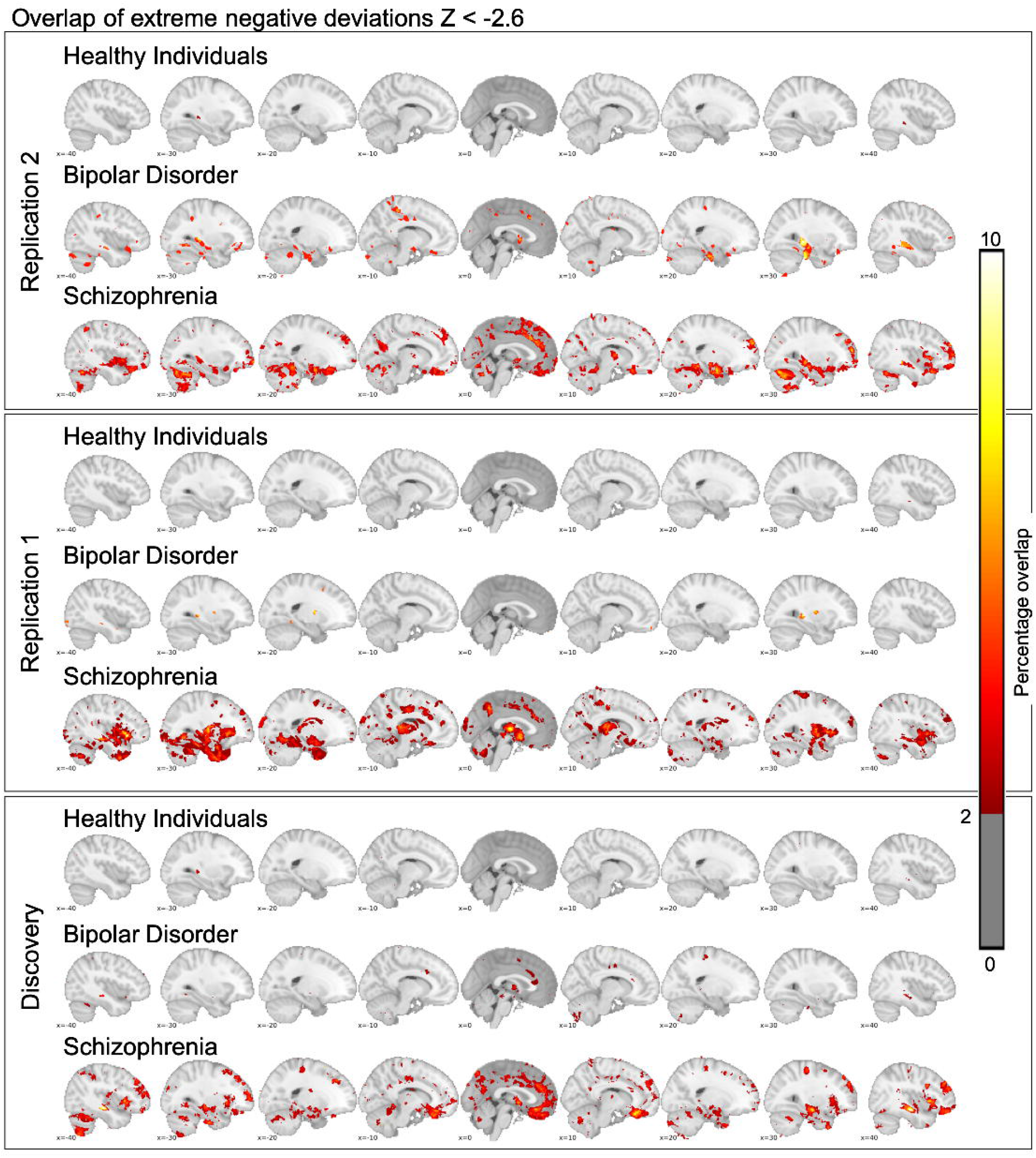
We show extreme negative deviations for healthy individuals, bipolar disorder and schizophrenia. In the lower panel we depict results based on the data reported in Wolfers et al. 2018, Jama Psychiatry. In the upper two panels we depict two replications. We show that the overlap across studies is comparable with only a few brain regions showing overlap in more than 2% of the individuals. While the spatial overlap is similar especially for schizophrenia there are also differences. Note that by comparing figure 2 and 3 it becomes apparent that robust group effects translate only to a relatively sparse overlap of extreme deviations from normality at the level of the individual. This replicates the main conclusion of the previous work. Note: Extreme negative deviations here are defined as Z < −2.6 at the individual level.

## DISCUSSION

Advanced brain imaging technology has allowed for probing the brain functional and structural correlates of complex human traits and mental disorders. While group-level normative deviations in brain structure in patients with a diagnosis of schizophrenia and bipolar disorder are robust and replicable (Figure 2) we observe substantial inter-individual differences in the neuroanatomical distribution of extreme deviations at the individual level (Figure 3, Supplementary Figure 4). These findings replicate and extend our previous study [Wolfers et al., 2018] and are largely in line with evidence of large heterogeneity across mental disorders [Wolfers et al., 2019; Zabihi et al., 2019].

Our results confirm that MRI-based brain structural aberrations in patients with severe mental disorders are highly heterogeneous in terms of their neuroanatomical distribution. These findings are in line with recent evidence of substantial brain structural heterogeneity in patients with schizophrenia [Alnæs et al., 2019] and also comply with accumulating evidence from psychiatric genetics strongly suggesting that mental illnesses are complex and heterogeneous disorders associated with a large number of genetic variants as well as environmental risk factors [Sullivan and Geschwind, 2019]. Along with documented clinical heterogeneity [Insel, 2009] and large interindividual variability in treatment response and outcome [Kapur et al., 2012], our successful replication of considerable neuroanatomical heterogeneity supports the need for statistical approaches that allow for inferences at the level of the individual. Characterizing the magnitude and distribution of brain aberrations in individual patients is key for identifying neuronal correlates of specific symptoms across diagnostic categories and would represent an important step towards increasing the utility of brain imaging in a clinical context.

While the present findings are robust, it must be considered that other data modalities beyond those provided by structural brain imaging may be more able to capture any common pathophysiological processes in patients with schizophrenia or bipolar disorder. Thus, we may observe larger overlaps across individuals with the same mental disorders in other data domains, such as those measuring brain function, cognition or specific behaviors, on the network-level or relevant biological assays. While this possibility cannot be ruled out the present results indicate that multiple pathophysiological processes and pathways are at play, which is also supported by the large number of identified genetic variants [Ripke et al., 2014; Smoller et al., 2013; Stahl et al., 2019].

Over the last decades it has become increasingly apparent that replication attempts in psychology, psychiatry, neuroscience and related fields frequently fail [Avinun et al., 2018; Dinga et al., 2019; Eklund et al., 2016; Hong et al., 2019; Ioannidis, 2005; Open Science Collaboration*, 2015; Tackett et al., 2019], which has fueled initiatives promoting reproducible science [Munafò et al., 2017; Poldrack et al., 2017; Schooler, 2014]. The neuroimaging field is no exception, and lack of reproducibility in brain imaging studies have been attributed to the high researcher degree of freedom in terms of the many and sometimes arbitrary analytical choices [Eklund et al., 2016]. Here we strictly adhered to the analytical protocols as specified in our original study [Wolfers et al., 2018]. The entire analytical pipeline is made publicly available to ease replication by independent researchers and to allow for application to different cohorts and disorders. Note here that if you replicate these findings in a sample on multiple scanners using different scanning sequences your interpretation might be misguided due to scanning confounds. Currently we are working on methods to improve normative modeling across sites[Kia et al., 2020]. While we are convinced that the here used analytical protocols are appropriate for testing the reproducibility of our original report, the approaches will be improved in future studies and are under continuous development [Kia et al., 2020; Kia and Marquand, 2018]. Moving forward, we will scale up this work towards larger samples covering a wider age range including neurodevelopment, incorporate different modalities and levels of information e.g. brain network level, including genetics, and link different experimental designs to the normative modeling framework.

Our replications support that group level-differences in brain structure disguise considerable individual differences in brain aberrations. While we find additional similarity across discovery and replication study (Supplementary Figure 2/3), such as extreme negative deviations are on average 3.91, 4.00 and 4.95 times more prevalent in individuals with schizophrenia than in healthy controls, we also find differences. Especially with respect to extreme positive deviations the pattern of overlap is as similar as it is different across studies (Supplementary Figure 4). However, when we look at the same pattern with a Z-threshold of 1.96 the overlap of extreme positive deviations shows striking similarities (Supplementary Figure 5-right panel). Further, we could not replicate a main group effect of bipolar disorder in comparison with healthy individuals in the cerebellum while this effect was present in the discovery sample (Figure 2). This may have been caused by differences in sample size. In addition, we want to point out that we worked on a predominantly adult sample, however, the onset of schizophrenia is primarily in adolescence or early adulthood. Therefore, it is important to investigate individual differences in this age group in future studies. Finally, we show results in addition to our previous study such as the correlation of extreme positive and negative deviations with PANSS scores. These results show that extreme negative deviations were associated with higher PANSS scores across all three samples but that this effect was only present when we pooled the bipolar and schizophrenia groups suggesting that it was driven by an increased power or by differences between the bipolar and schizophrenia patients rather than higher symptom scores. This interpretation is in line with the fact that we could replicate all previous findings of extreme negative deviations from normality across the two replication samples (Table 2). With low reproducibility rates across various scientific disciplines [Baker and Penny, 2016] building confidence through replication is critical.

## CONCLUSION

Individuals with a mental disorder are sampled from a heterogenous general population based on their clinical and symptom profiles. One would expect a higher degree of similarity in terms of normative deviations in patients with the same diagnosis than in healthy individuals on measures affected by the disorder. In other words, these deviations would be enriched in the clinical as opposed to the general population. This is exactly what we observe and replicate across three samples. However, we do not detect it to the degree that group studies would suggest which generally show significant differences between patients and healthy individuals in terms of averages. Consequentially, these group differences say little about the individual patient with a mental disorder and his/her deviation from a norm, pointing out that we need individualized analyses instead of a focus on group studies in psychiatry. Therefore, the main conclusion of the discovery study is supported by replications across two samples, namely that group level-differences in brain structure captured by a classical case-control paradigm, disguises considerable individual differences in brain aberrations when we map deviations from normality.

## Data Availability

As the data is clinical data including patient information access is restricted in accordance with European and Norwegian law.

## ACKNOWLEDGEMENTS

This work was performed on the TSD (Tjenester for Sensitive Data) facilities, owned by the University of Oslo, operated and developed by the TSD service group at the University of Oslo, IT-Department (USIT) with resources provided by UNINETT Sigma2 - the National Infrastructure for High Performance Computing and Data Storage in Norway. The study is supported by the Research Council of Norway (223273, 249795, 298646, 300768, 276082), the South-Eastern Norway Regional Health Authority (2014097, 2015073, 2016083, 2019101), a Wellcome Trust Innovator award (‘BRAINCHART’, 215698/Z/19/Z), and the European Research Council under the European Union’s Horizon 2020 research and Innovation program (ERC StG Grant 802998 and Grant 847776). TW gratefully acknowledges the Niels Stensen Fellowship as well as the European Union’s Horizon 2020 research and innovation programme under the Marie Sklodowska-Curie grant agreement No 895011. Data is available upon on request due to privacy/ ethical restrictions. We would like to thank the participants of these studies for their contribution.

