## Supplement for "Replicating extensive brain structural heterogeneity in individuals with schizophrenia and bipolar disorder" (from another version of this preprint: version 2, 2021-01-13)

### SUPPLEMENTAL MATERIALS

**Supplementary Table 1: Replication of extreme deviations with different thresholds**

**Extreme deviations (|Z|>1.96)**

| | Replication 2 | | | Replication 1 | | | Discovery | | |
|---|---|---|---|---|---|---|---|---|---|
| ***Case-Control*** | **Healthy** | **BP** | **SZ** | **Healthy** | **BP** | **SZ** | **Healthy** | **BP** | **SZ** |
| *Extreme negative (mean, std)* | 1.71 +- 2.72% | 2.09+- 4.08% | 4.34+- 7.97% | 1.50 +- 2.44 | 1.44+- 2.01% | 4.03+- 4.70% | 1.62 +- 2.82% | 2.00 +- 2.71% | 4.87 +- 6.38% |
| *Significance* | HC = BD<br>**HC < SZ (p < 0.001)**<br>**BP < SZ (p < 0.005)** | | | HC = BD<br>**HC < SZ (p < 0.001)**<br>**BP < SZ (p < 0.001)** | | | HC = BD<br>**HC < SZ (p < 0.001)**<br>**BP < SZ (p < 0.001)** | | |
| *Extreme positive (mean, std)* | 3.39 +- 4.56% | 3.06 +- 4.93% | 2.74 +- 3.90% | 3.59 +- 4.65% | 2.87 +- 3.68% | 2.06 +- 2.44% | 3.47 +- 4.29% | 2.73 +- 3.35% | 2.40 +- 3.87% |
| *Significance* | HC = BD<br>**HC > SZ (p < 0.05)**<br>BP = SZ | | | HC = BD<br>**HC > SZ (p < 0.001)**<br>**BP > SZ (p < 0.05)** | | | HC = BD<br>**HC > SZ (p < 0.001)**<br>**BP > SZ (p < 0.01)** | | |

**Extreme deviations (|Z|>3.1)**

| | Replication 2 | | | Replication 1 | | | Discovery | | |
|---|---|---|---|---|---|---|---|---|---|
| | **Healthy** | **BP** | **SZ** | **Healthy** | **BP** | **SZ** | **Healthy** | **BP** | **SZ** |
| *Extreme negative (mean, std)* | 0.04 +- 0.18% | 0.07+- 0.27% | 0.39+- 1.95% | 0.02 +- 0.07% | 0.01+- 0.06% | 0.15+- 0.62% | 0.06 +- 0.37% | 0.03+- 0.10% | 0.20+- 0.81% |
| *Significance* | HC = BD<br>**HC < SZ (p < 0.001)**<br>**BP < SZ (p < 0.05)** | | | HC = BD<br>**HC < SZ (p < 0.001)**<br>**BP < SZ (p < 0.001)** | | | HC = BD<br>**HC < SZ (p < 0.001)**<br>**BP < SZ (p < 0.001)** | | |
| *Extreme positive (mean, std)* | 0.39 +- 1.09% | 0.40+- 1.47% | 0.32+- 0.74% | 0.46 +- 0.93 | 0.33+- 0.62% | 0.23+- 0.43% | 0.41 +- 0.81% | 0.28+- 0.53% | 0.30+- 0.66% |
| *Significance* | HC = BD<br>HC = SZ<br>BP = SZ | | | HC = BD<br>**HC > SZ (p < 0.001)**<br>**BP > SZ (p < 0.05)** | | | HC = BD<br>**HC > SZ (p < 0.005)**<br>**BP > SZ (p < 0.05)** | | |

**Extreme deviations (EVD)**

| | Replication 2 | | | Replication 1 | | | Discovery | | |
|---|---|---|---|---|---|---|---|---|---|
| | **Healthy** | **BP** | **SZ** | **Healthy** | **BP** | **SZ** | **Healthy** | **BP** | **SZ** |
| *Extreme values (mean, std)* | 0.46 +- 0.28 | 0.45+- 0.27 | 0.61+- 0.25 | 0.48 +- 0.28 | 0.43+- 0.28 | 0.61+- 0.28 | 0.43 +- 0.27 | 0.45+- 0.27 | 0.62+- 0.27 |
| *Significance* | HC = BD<br>**HC < SZ (p < 0.001)**<br>**BP < SZ (p < 0.001)** | | | HC = BD<br>**HC < SZ (p < 0.001)**<br>**BP < SZ (p < 0.001)** | | | HC = BD<br>**HC < SZ (p < 0.001)**<br>**BP < SZ (p < 0.001)** | | |

*NOTE: PANSS, Positive and Negative Syndrome Scale.*

**Supplementary Table 2: Correlations of extreme deviations |Z|>2.6 with symptom scores**

| ***Across Groups*** | | **Replication 2** | **Replication 1** | **Discovery** |
|---|---|---|---|---|
| ***Extreme deviations*** | ***Symptoms**** | **BP & SZ** | **BP & SZ** | **BP & SZ** |
| *Extreme negative deviations* | *PANSS global* | r=0.149<br>p>0.05 | r=0.033<br>p>0.05 | **r=0.179**<br>**p<0.001** |
| | *PANSS negative* | **r=0.232**<br>**p<0.005** | **r=0.190**<br>**p<0.01** | **r=0.217**<br>**p<0.001** |
| | *PANSS positive* | r=0.101<br>p>0.05 | **r=0.198**<br>**p<0.005** | **r=0.192**<br>**p<0.001** |
| | *PANSS total* | **r=0.190**<br>**p<0.05** | **r=0.157**<br>**p<0.05** | **r=0.241**<br>**p<0.001** |
| ***Extreme deviations*** | ***Symptoms**** | **BP&SZ** | **BP&SZ** | **BP&SZ** |
| *Extreme positive deviations* | *PANSS global* | r=0.011<br>p>0.05 | r=-0.094<br>p>0.05 | r=0.045<br>p>0.05 |
| | *PANSS negative* | r=0.073<br>p>0.05 | r=-0.061<br>p>0.05 | r=0.049<br>p>0.05 |
| | *PANSS positive* | r=0.124<br>p>0.05 | r=-0.056<br>p>0.05 | r=0.019<br>p>0.05 |
| | *PANSS total* | r=0.071<br>p>0.05 | r=-0.072<br>p>0.05 | r=0.035<br>p>0.05 |

| **Within Groups** | | **Replication 2** | | **Replication 1** | | **Discovery** | |
|---|---|---|---|---|---|---|---|
| ***Extreme deviations*** | ***Symptoms**** | **BP** | **SZ** | **BP** | **SZ** | **BP** | **SZ** |
| *Extreme negative deviations* | *PANSS global* | r=0.047<br>p>0.05 | r=0.078<br>p>0.05 | r=0.106<br>p>0.05 | r=-0.082<br>p>0.05 | r=0.060<br>p>0.05 | r=0.104<br>p>0.05 |
| | *PANSS negative* | r=0.068<br>p>0.05 | r=0.183<br>p>0.05 | r=0.091<br>p>0.05 | r=-0.052<br>p>0.05 | r=-0.002<br>p>0.05 | r=0.154<br>p>0.05 |
| | *PANSS positive* | r=-0.044<br>p>0.05 | r=0.017<br>p>0.05 | r=0.045<br>p>0.05 | r=-0.021<br>p>0.05 | r=0.014<br>p>0.05 | r=0.091<br>p>0.05 |
| | *PANSS total* | r=0.063<br>p>0.05 | r=0.121<br>p>0.05 | r=-0.008<br>p>0.05 | r=-0.068<br>p>0.05 | r=0.056<br>p>0.05 | r=0.132<br>p>0.05 |
| ***Extreme deviations*** | ***Symptoms**** | **BP** | **SZ** | **BP** | **SZ** | **BP** | **SZ** |
| *Extreme positive deviations* | *PANSS global* | r=-0.040<br>p>0.05 | r=0.051<br>p>0.05 | r=-0.133<br>p>0.05 | r=0.038<br>p>0.05 | **r=0.186**<br>**p<0.05** | r=0.012<br>p>0.05 |
| | *PANSS negative* | r=-0.072<br>p>0.05 | r=0.174<br>p>0.05 | r=-0.082<br>p>0.05 | r=0.120<br>p>0.05 | **r=0.212**<br>**p<0.05** | r=0.073<br>p>0.05 |
| | *PANSS positive* | r=-0.052<br>p>0.05 | **r=0.244**<br>**p<0.05** | r=-0.029<br>p>0.05 | r=0.068<br>p>0.05 | **r=0.175**<br>**p<0.05** | r=-0.005<br>p>0.05 |
| | *PANSS total* | r=-0.069<br>p>0.05 | r=0.150<br>p>0.05 | r=-0.092,<br>p>0.05 | r=0.090<br>p>0.05 | **r=0.215**<br>**p<0.05** | r=0.032<br>p>0.05 |

*Abbreviations: PANSS, Positive and Negative Syndrome Scale. BP, bipolar disorder. SZ, schizophrenia.*

* Symptom scores have been assessed using PANSS which is a standard clinical instrument for the quantification of positive and negative psychotic symptoms.

*Supplementary Figure 1: In the upper panels we depict the mean Z-scores based on the PALM results for individuals that are healthy or those that hold a diagnosis of bipolar disorder or schizophrenia for the two replication samples. In the lower panel we depict results based on the data reported in Wolfers et al. 2018, Jama Psychiatry.*

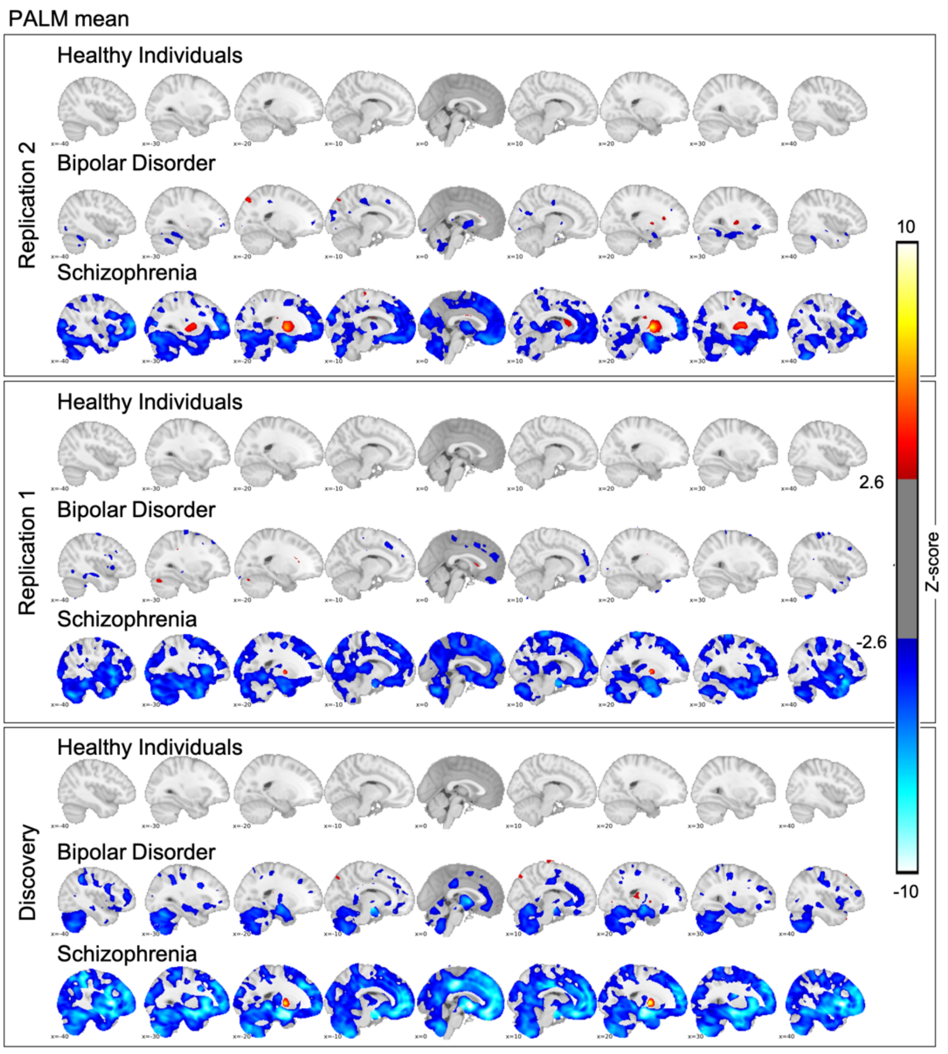

*Supplementary Figure 2: Percentage extreme deviations at |Z|=2.6 per participants plotted as boxplot for each group. The results replicate well across samples. In the lower panel we depict results based on the data reported in Wolfers et al. 2018, Jama Psychiatry.*

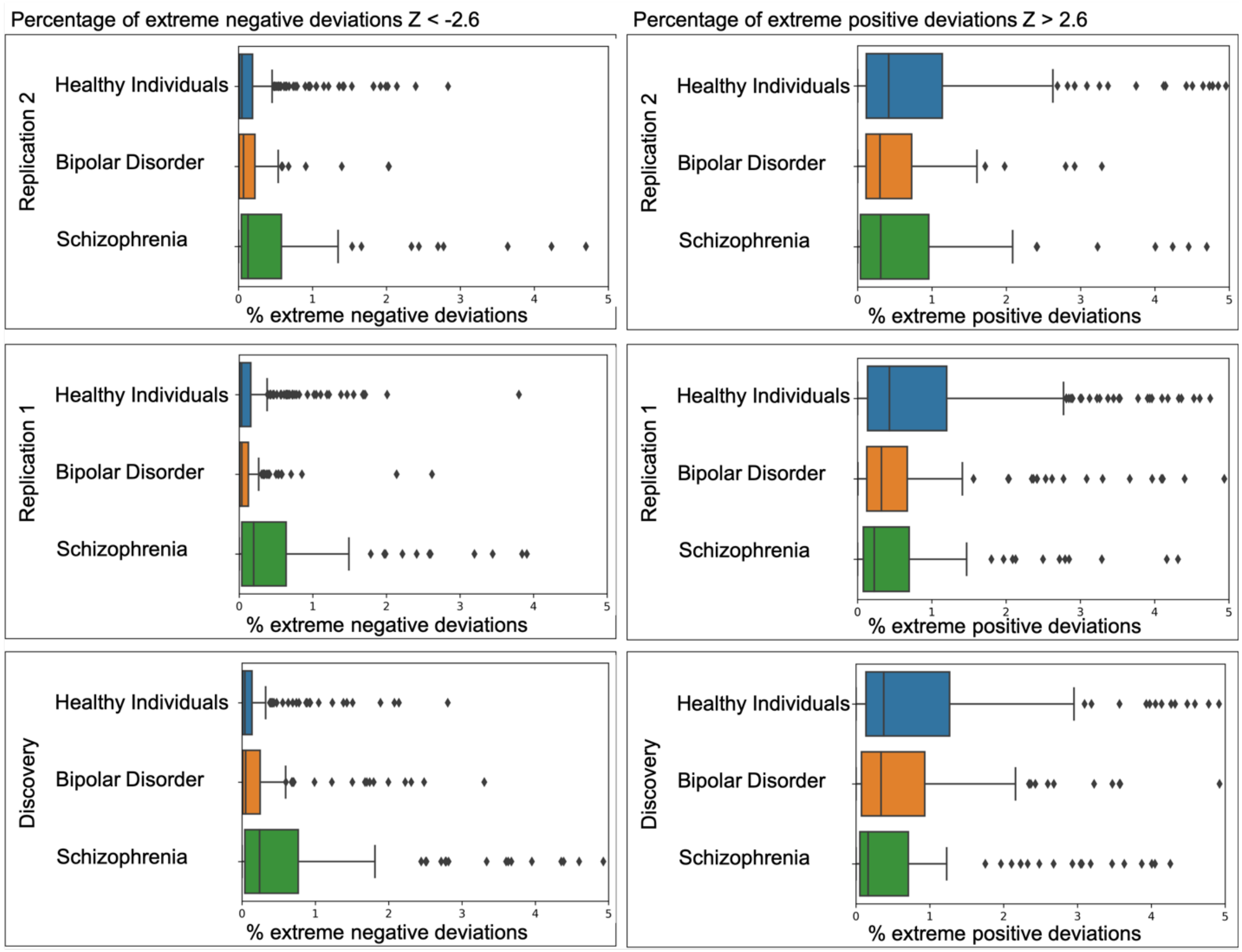

*Supplementary Figure 3: Percentage extreme deviations at |Z|=2.6 per participants plotted as violin-plot for each group. The results replicate well across samples. In the lower panel we depict results based on the data reported in Wolfers et al. 2018, Jama Psychiatry.*

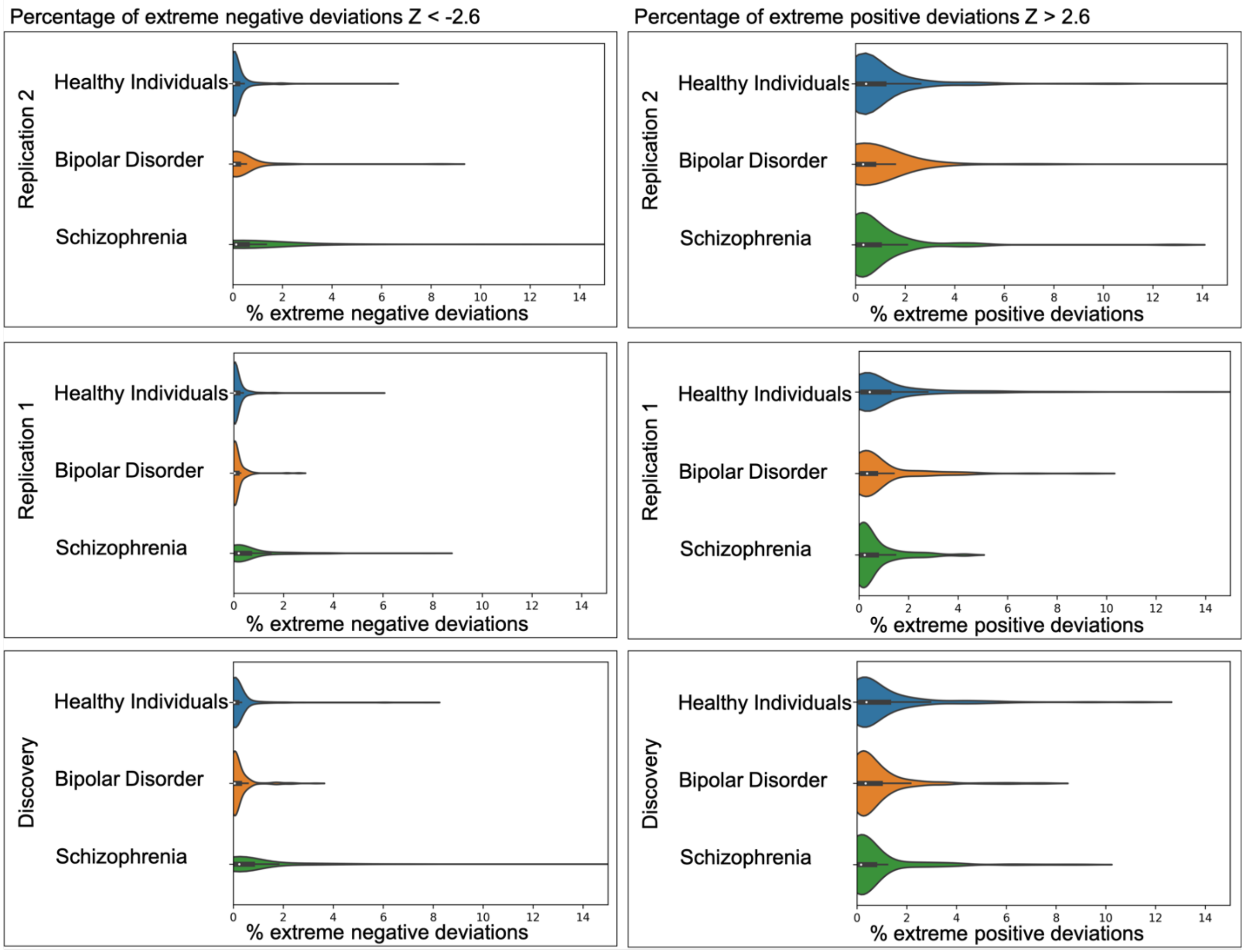

*Supplementary Figure 4: Overlap of extreme positive deviations from normality for healthy individuals, individuals with bipolar disorder and schizophrenia. The different samples show comparable results with only a few brain regions showing overlap in more than 2% of the individuals diagnosed with the same mental disorder. In replication 1 peak voxels show extreme positive overlap in 4.66% in healthy individuals, 8.15% in individuals with bipolar disorder and 6.13% in schizophrenia. In replication 2 peak voxels show extreme positive overlap in 4.48% in healthy individuals, 11.47% in individuals with bipolar disorder and 10.47% in schizophrenia. In the lower panel we depict results based on the data reported in Wolfers et al. 2018, Jama Psychiatry. Note: This overlap is based on a Z-threshold of larger than 2.6.*

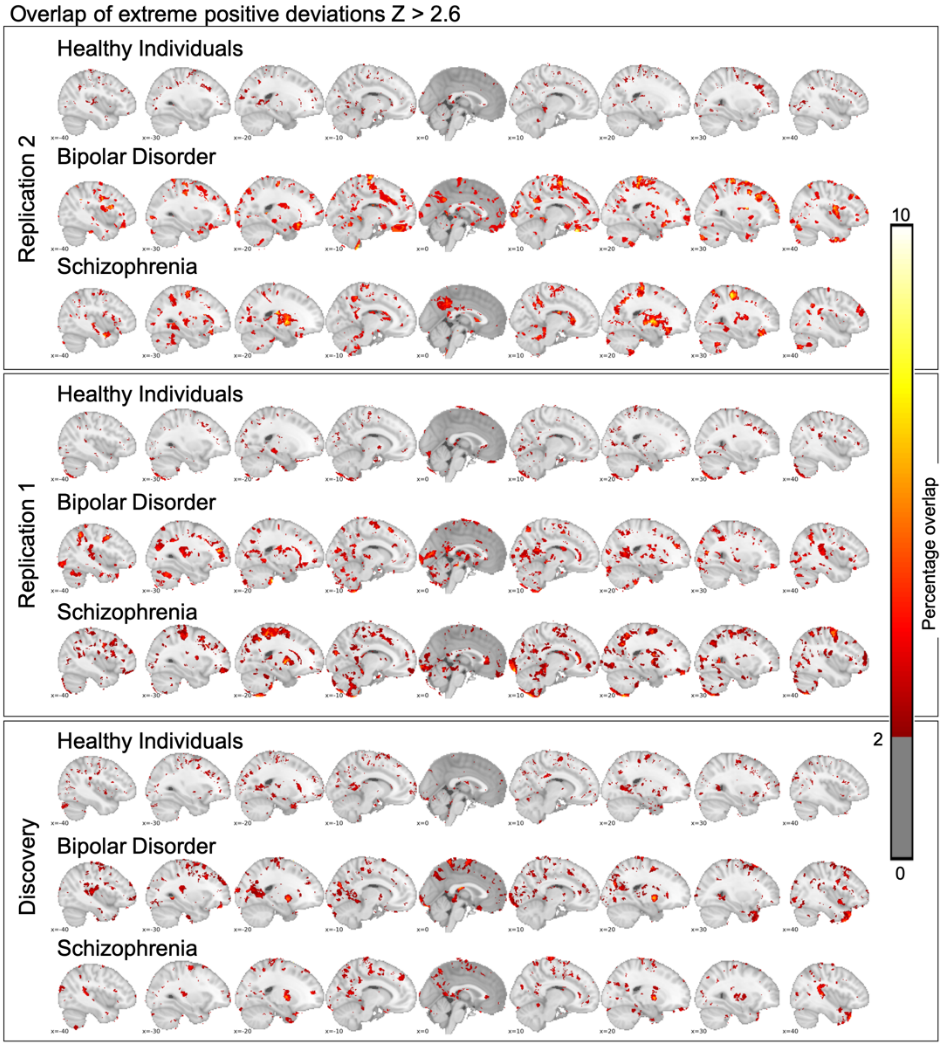

*Supplementary Figure 5: Overlap of extreme negative and positive deviations from normality for healthy individuals, individuals with bipolar disorder and schizophrenia with a normative probability map thresholded at |Z|=1.96. In expectations a lower threshold yields higher overlap, however, the overlap remains small with only a few brain regions showing extreme negative deviations in more than 5% of the patients (yellow regions). Extreme positive deviations show a widespread overlap in healthy individuals when thresholded at 2%. However, taking a threshold of 5% no region overlaps in healthy individuals (yellow regions). Both extreme negative and positive deviations show peak overlap in the same regions as in Figure 3 and Supplementary Figure 2. In the lower panel we depict results based on the data reported in Wolfers et al. 2018, Jama Psychiatry, which show striking similarity with the results obtained from both replication samples.*

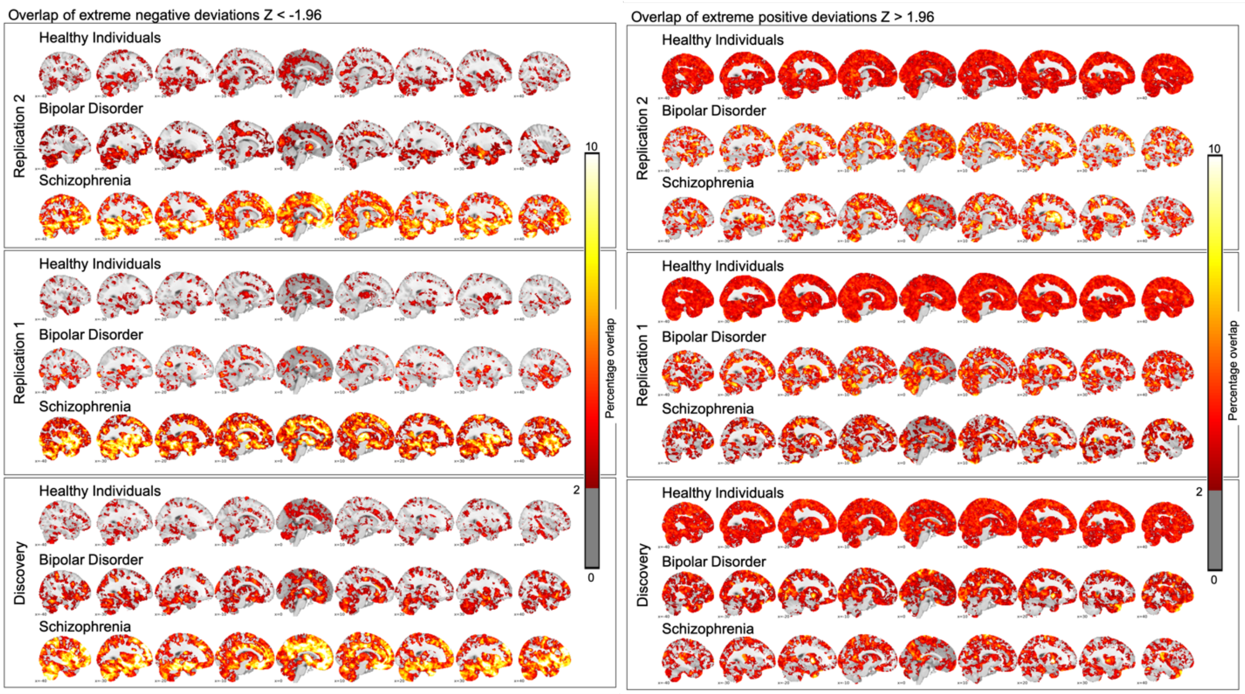

*Supplementary Figure 6: Overlap of extreme negative and positive deviations from normality for healthy individuals, individuals with bipolar disorder and schizophrenia with a normative probability map thresholded at |Z|=3.1. In expectations a higher threshold yields lower overlap. Therefore, we observe less overlap in extreme positive and extreme negative deviations from normality. These overlaps are widespread and peak in some of the regions as in Figure 3 and Supplementary Figure 2. In the lower panel we depict results based on the data reported in Wolfers et al. 2018, Jama Psychiatry.*

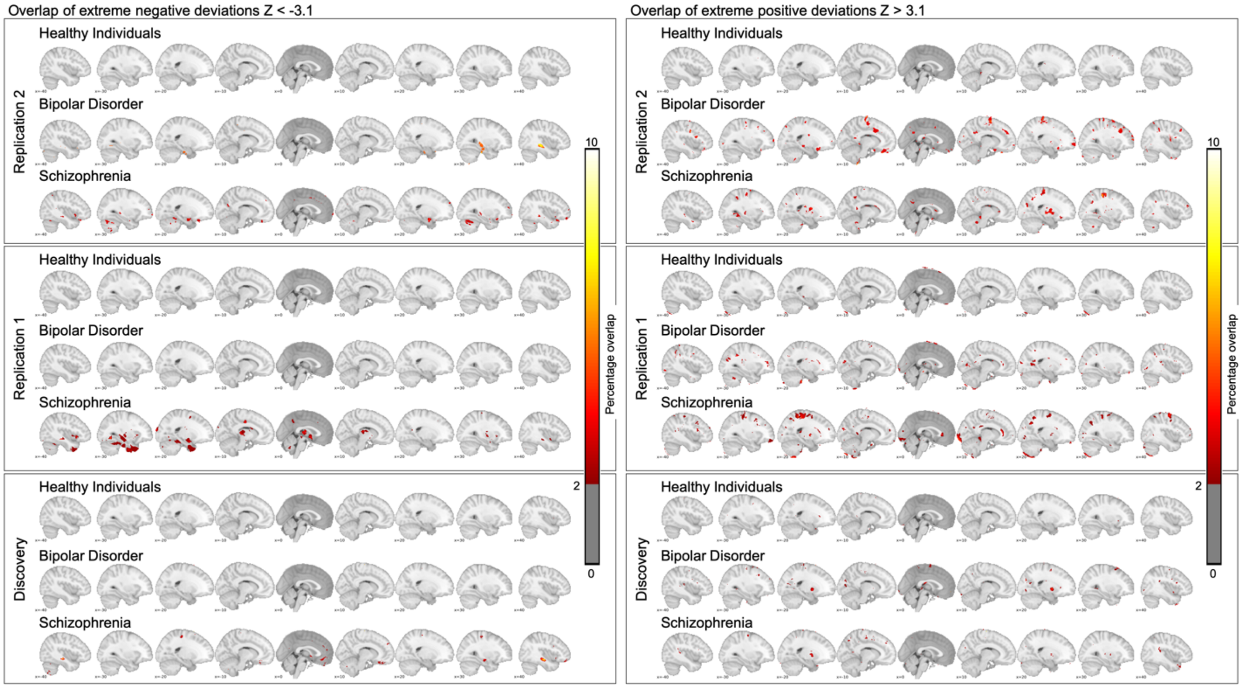

*Supplementary Figure 7: Overlap of extreme negative and positive deviations from normality for healthy individuals, individuals with bipolar disorder and schizophrenia with a normative probability map thresholded at FDR=0.05. In expectations a higher threshold yields lower overlap. Therefore, we observe less overlap in extreme positive and extreme negative deviations from normality which are comparable to |Z|>3.1. These overlaps are widespread and peak in some of the regions as in Figure 3 and Supplementary Figure 2. In the lower panel we depict results based on the data reported in Wolfers et al. 2018, Jama Psychiatry.*

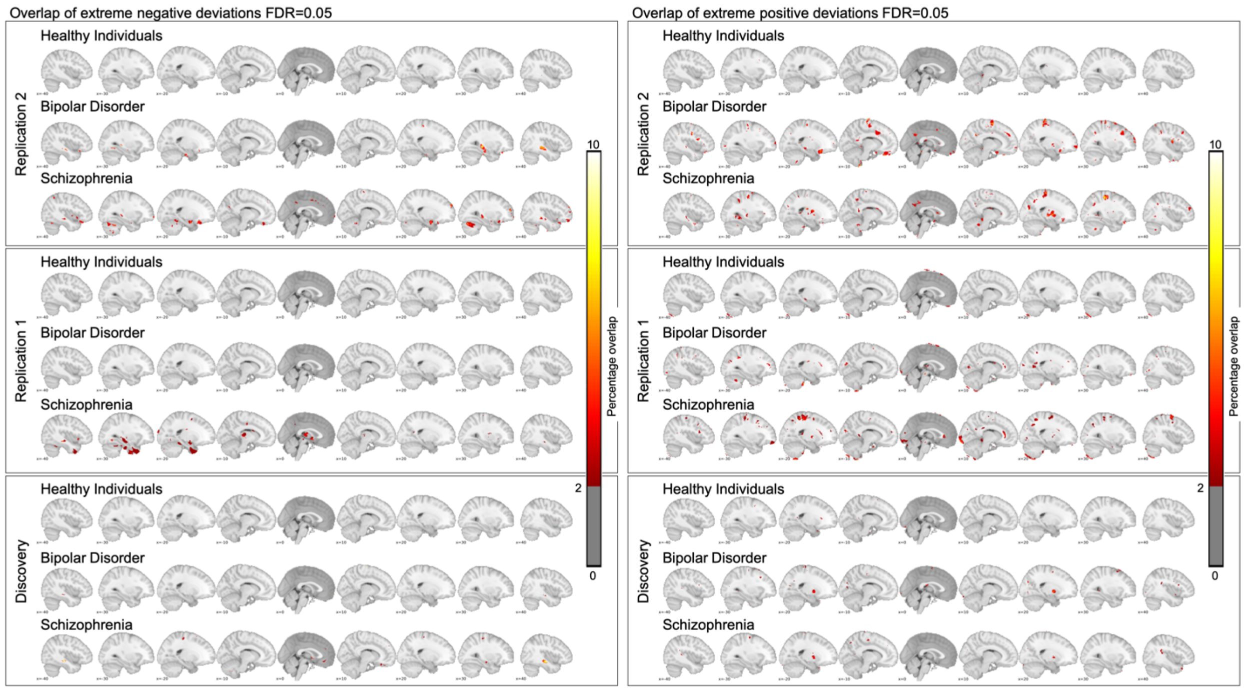

*Supplementary Figure 8: Performance evaluation of the normative models under 10-fold cross-validation. Reported are the correlations between predicted and observed values without threshold, with threshold of |r|>0.4 and the corresponding 1-p-values thresholded at 1-p>.095. It is apparent that the normative models were predictive across the whole brain across three samples. In the lower panel we depict results based on the data reported in Wolfers et al. 2018, Jama Psychiatry.*

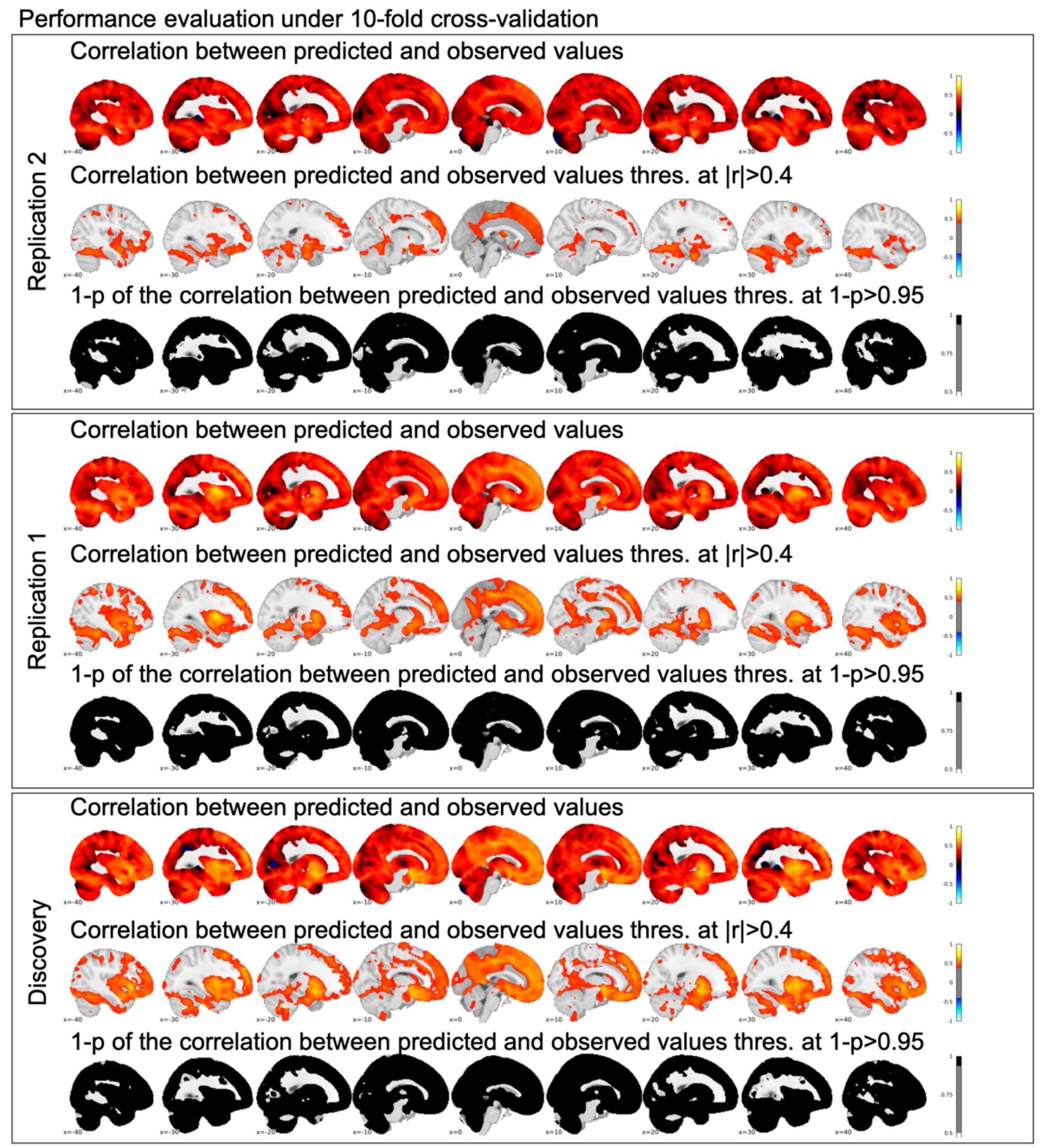
